# The National Transportation Noise Exposure Map

**DOI:** 10.1101/2023.02.02.23285396

**Authors:** Edmund Seto, Ching-Hsuan Huang

**Author notes:** Corresponding Author: Edmund Seto, Associate Professor, Department of Environmental & Occupational Health Sciences, University of Washington, 4225 Roosevelt Way NE, Suite 100, Seattle, WA 98105, United States.

## Abstract

We describe the development and public availability of the National Transportation Noise Exposure Map with the goal of estimating population exposures to various noise levels at the census tract level in the United States. The map was created by overlaying the Bureau of Transportation Statistics’ National Transportation Noise Map with 5-year block group population estimates from the American Community Survey, and aggregating exposed population estimates to the census tract level. Based on the exposure map, an estimated 94.9 million people (29.1 % of the total U. S. population) were exposed to ≥ 45 dB LAeq of transportation-related noise, and approximately 11.9 million (3.6 %) were exposed to ≥ 60 dB in the year 2020.The exposure maps indicate that the greatest population proportion and number of individuals exposed were in California, while generally the map illustrates high proportions of exposure for populations living along major U. S. roadways and in airport communities. The availability of this new exposure map will facilitate the integration of noise exposures into a variety of studies, including regional and national health impact assessments, epidemiologic, and environmental justice studies.

## Introduction

Community noise has been linked to numerous negative health effects, including high annoyance (1,2), poor mental health (3), obesity (4), hypertension (5), heart disease (6–8), increased risk of diabetes (9), and sleep disturbance (10,11). Environmental noise affects some population groups more than others (12). Children exposed to high levels of noise experienced increased adrenaline and noradrenaline levels, annoyance, hyperactivity, high blood pressure, and poorer performance related to reading comprehension and memory skills and standardized academic tests (13). Noise exposures among the elderly have been found to be associated with hypertension (14) and stoke (15). Assessment of population exposures to environmental noise is therefore important for understanding its role in the health of communities.

Mapping of environmental noise levels and assessment of population exposures is required of member states under the European Union’s Environmental Noise Directive (END) (16). These maps and exposure estimates inform the EU’s health impact assessments (17). Yet, nationwide population noise exposure mapping is typically not conducted for the United States due to the lack of city, state, and federal mandates and funding (18). Previous U. S. studies have largely focused on mapping noise exposures and health impacts for individual cities and regions (19– 21). However, the U. S. EPA conducted a nationwide noise assessment in1981, which estimated population exposures to various noise level categories (22). Although the EPA’s assessment did not result in spatially explicit population exposures similar to those conducted under the END, the overall estimates of population numbers exposed to various noise level categories would enable use of noise exposure-response relationships to estimate various population health impacts, similar to how noise health impact assessments are conducted in the EU. More recent and promising efforts to map noise levels and exposures in the continental U. S., include a spatial model developed from noise monitoring data collected at various urban and rural sites, and principally at sites in national parks as required by the U. S. National Park Service (NPS) (23–26).

The U. S. Bureau of Transportation Statistics (BTS) developed the National Transportation Noise Map (NTNM), a detailed modeled noise map that covers the continental U. S., as well as the states of Alaska and Hawaii (27). The NTNM is intended to serve as a transportation planning tool, and therefore is based on modeled noise estimates from certain transportation sources, such as roadway, aviation, and rail traffic. While the BTS mention that the noise map data allow for viewing of potential exposures, the map itself only shows sound levels. While it is possible to overlay the NTNM with population data from the census to quantify and map population numbers exposed different transportation noise level categories, thus far a national transportation noise exposure map is not widely available. A regularly updated and easily accessed U. S. noise exposure map would greatly facilitate health impact assessments, health effects studies that wish to incorporate estimated transportation-related noise exposure, and environmental justice studies.

In this paper we describe the development and public availability of national transportation exposure map for the U. S. with the goal of estimating population exposures to various noise levels at the census tract level. This map is based on an overlay of the most recently available NTNM and spatial population estimates from the American Community Survey (ACS) first at the census block group level for improved spatial accuracy, which is then aggregated to the census tract level for easier application to various population health studies. Based on this map, we present summaries of population numbers and proportions exposed to various noise levels at the state and national level. In the Discussion, we compare our findings with the EPA’s 1981 noise assessment, and compare our spatial analysis approach to the modeling based on other recent noise modeling work for the U.S. We describe the potential limitations of the NTNM and our exposure map for those intending to use the exposure information for health impact assessment, epidemiologic, and environmental justice studies.

## Methods

Spatial raster files of noise levels related to aviation, roadway and rail traffic were obtained from the BTS (28). These files were separated by state, with the most recent results available for the year 2020. The modeled noise levels in the NTNM represent potential noise levels on an average day within the year 2020. The noise metric is the LAeq, the 24-hour equivalent A-weighted sound level at modeled receptor locations. Noise levels below 45 dBA LAeq are not included in the raster files, and pixels below this threshold are assigned missing values. The estimated noise levels at receptor locations from separate aviation, road, and rail models are acoustically summed to create a composite LAeq from the combined noise sources. We briefly describe the aviation, roadway, and trail models used below. Modeling details and assumptions are provided by the BTS (28).

Noise attributable to aviation was modeled in the NTNM based on the Federal Aviation Administration (FAA) Aviation Environmental Design Tool (AEDT) model (29), using input from flight operation schedules and air traffic counts. Flight operations are averaged into a single typical average annual data, and include airports with an average of at least one daily flight departure. The input does not include airports that only have military operations, and does not include helicopter flights. The AEDT modeling approach uses a dynamic receptor grid spacing, in which an initial tenth of a mile grid around each airport is refined to a smaller spacing to produce a smooth noise contour is produced.

Noise attributable to roadway traffic was modeled in the Federal Highway Administration (FHWA) Traffic Noise Model (TNM) (30,31). TNM uses roadway traffic data in the form of average annual daily traffic (AADT) traffic counts from the FHWA’s Highway Performance Monitoring System (HPMS) for various road types, including interstate highways, major and minor arterials, and major collector streets in both urban and rural areas. The traffic counts used by the model include automobiles and medium and heavy trucks. TNM is calculated for a 30 m grid of receptor locations.

Rail noise from conventional passenger trains is based on the Federal Transit Administration (FTA) Transit Noise and Vibration Impact Assessment Manual (32), while noise from high-speed rail is based on the Federal Railroad Administration (FRA) High Speed Ground Transportation Noise and Vibration Impact Assessment Manual (33). The models use train schedule and traffic data that is meant to be representative of a typical weekday. The year 2020 NTNM also includes freight train traffic in the model. Rail noise is calculated for a 30 m receptor grid.

Population exposures to noise was estimated by overlaying the NTNM noise raster map and census block group 5-year (2016-2020) estimates of total population from the American Community Survey (ACS) obtained through the Census Bureau API (34). The proportion of each census block group area that overlapped with LAeq noise categories of 45-50, 50-60, 60-70, 70-80, 80-90 and 90+ dB (greater than or equal applied to the lower bound of each category, and less than applied to the upper bound). This proportion was assumed to be the same as the proportion of the block group population exposed to the respective noise level category. The proportion of area in each block group with missing values for noise exposure was assumed to be equivalent to the proportion of the block group population that was exposed to less than LAeq 45 dB of transportation-related noise. For easier merging with census demographic information in future studies, the block group exposure estimates were aggregated to the census tract level. This results in what we call the National Transportation Noise Exposure Map (NTNE Map).

Data processing and analyses were conducted in R (version 4.2.2) and the stars (0.6.0), sf (1.0.9), exactextractr (0.8.2), ggplot2 (3.4.0), and dplyr (1.0.10) packages. Data are provided as R sf data objects and as shapefiles at the link below. An online interactive version of the NTNE Map that allows for zooming and address/geolocation search is available at: https://deohs.washington.edu/national-transportation-noise-exposure-map

## Results

Population numbers and proportions exposed to each of the noise level categories are presented by state and for the nation in Table 1. An estimated 94.9 million people (29.1 % of the total population) were exposed to ≥ 45 dB LAeq of transportation-related noise, and approximately 11.9 million (3.6 %) were exposed to ≥ 60 dB in the U.S. in 2020. The census tract-level map of the proportion exposed to LAeq ≥ 60 dB for the continental US is shown in Figure 1 (maps of Alaska and Hawaii are provided in Supplemental Info as Figures S1 and S2).

**Table 1.**
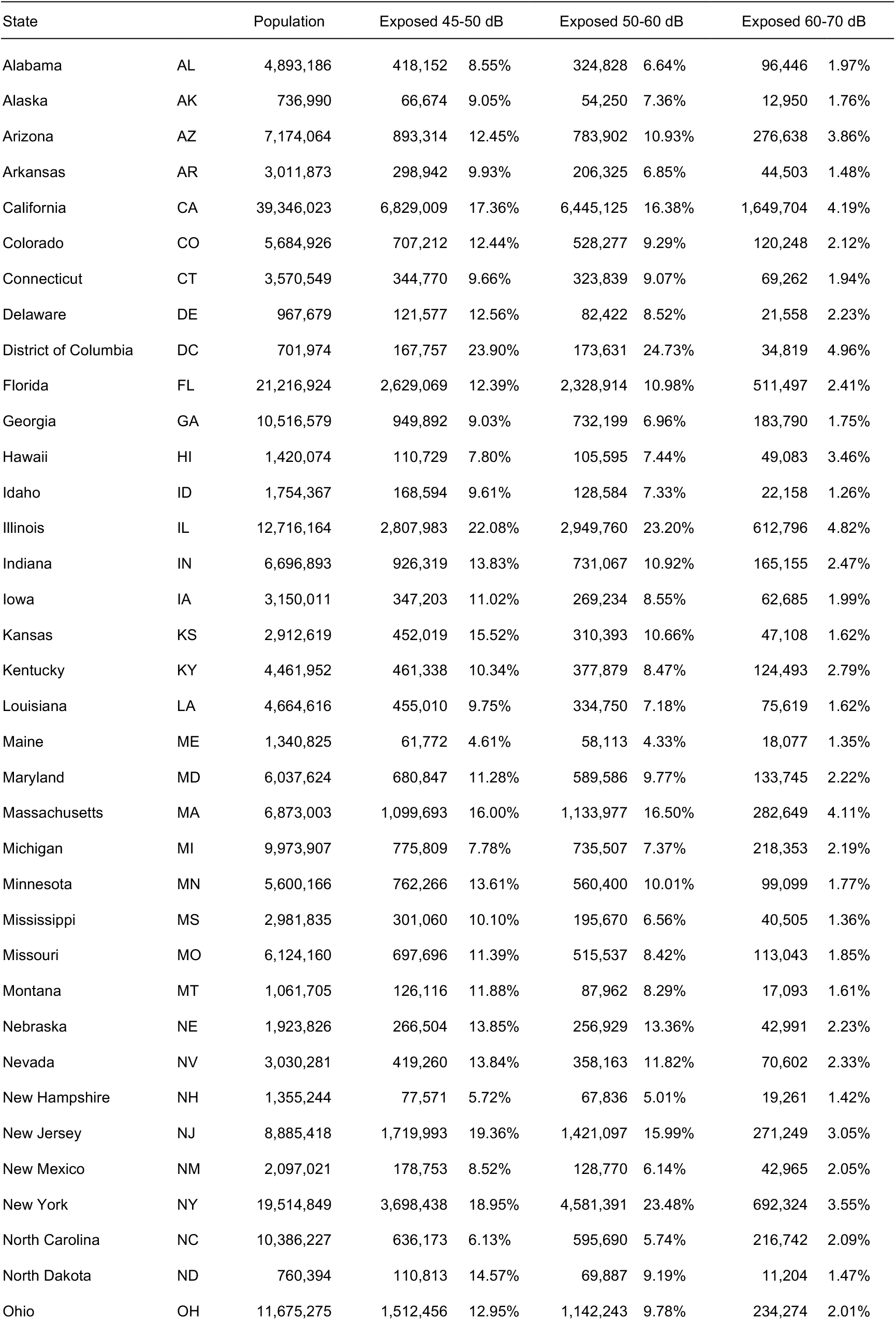
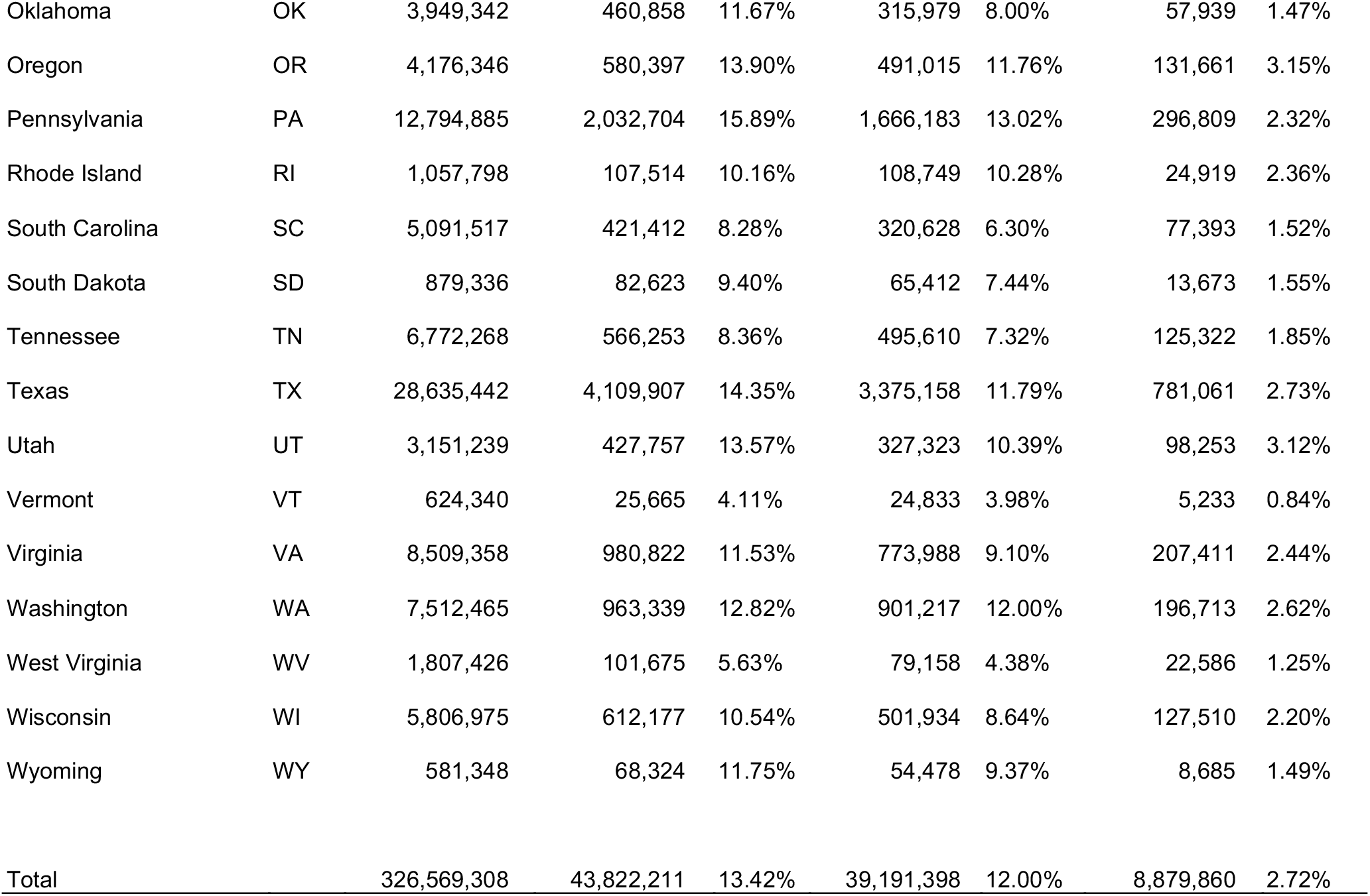
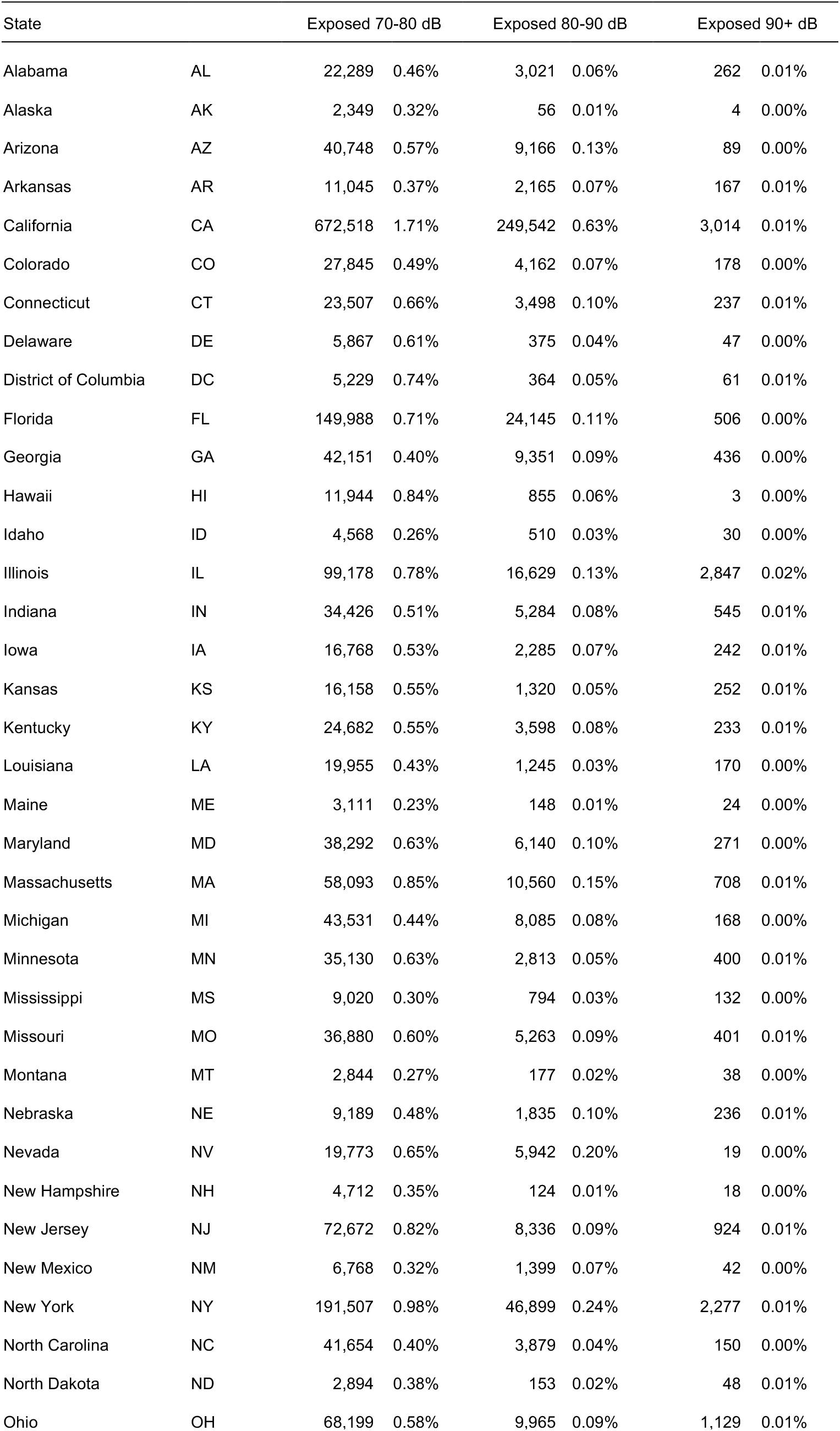
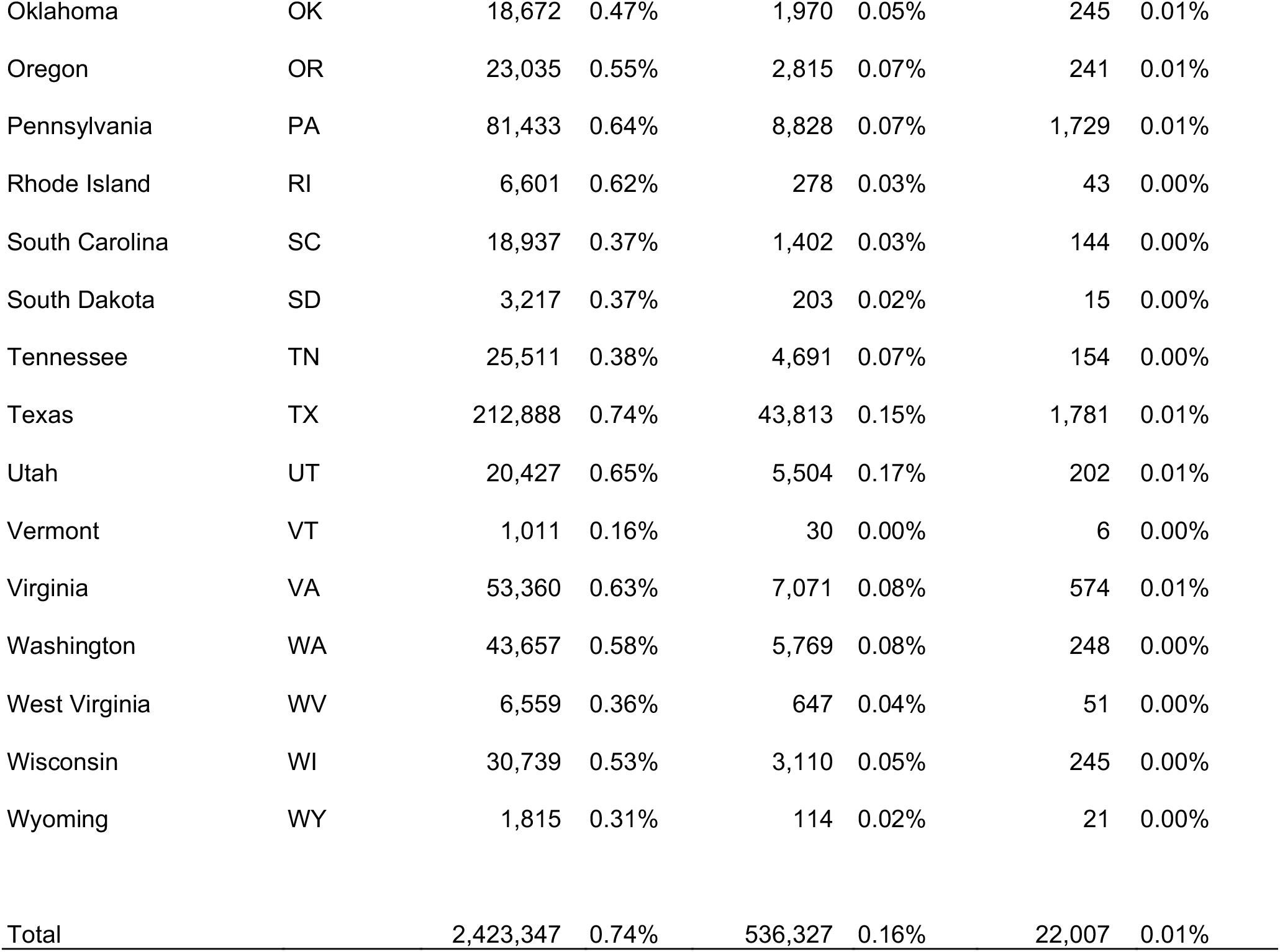
Population numbers and proportions exposed to noise levels by state and total for all the United States.

| State |  | Population | Exposed 45-50 dB |  | Exposed 50-60 dB |  | Exposed 60-70 dB |  |
| --- | --- | --- | --- | --- | --- | --- | --- | --- |
| Alabama | AL | 4,893,186 | 418,152 | 8.55% | 324,828 | 6.64% | 96,446 | 1.97% |
| Alaska | AK | 736,990 | 66,674 | 9.05% | 54,250 | 7.36% | 12,950 | 1.76% |
| Arizona | AZ | 7,174,064 | 893,314 | 12.45% | 783,902 | 10.93% | 276,638 | 3.86% |
| Arkansas | AR | 3,011,873 | 298,942 | 9.93% | 206,325 | 6.85% | 44,503 | 1.48% |
| California | CA | 39,346,023 | 6,829,009 | 17.36% | 6,445,125 | 16.38% | 1,649,704 | 4.19% |
| Colorado | CO | 5,684,926 | 707,212 | 12.44% | 528,277 | 9.29% | 120,248 | 2.12% |
| Connecticut | CT | 3,570,549 | 344,770 | 9.66% | 323,839 | 9.07% | 69,262 | 1.94% |
| Delaware | DE | 967,679 | 121,577 | 12.56% | 82,422 | 8.52% | 21,558 | 2.23% |
| District of Columbia | DC | 701,974 | 167,757 | 23.90% | 173,631 | 24.73% | 34,819 | 4.96% |
| Florida | FL | 21,216,924 | 2,629,069 | 12.39% | 2,328,914 | 10.98% | 511,497 | 2.41% |
| Georgia | GA | 10,516,579 | 949,892 | 9.03% | 732,199 | 6.96% | 183,790 | 1.75% |
| Hawaii | HI | 1,420,074 | 110,729 | 7.80% | 105,595 | 7.44% | 49,083 | 3.46% |
| Idaho | ID | 1,754,367 | 168,594 | 9.61% | 128,584 | 7.33% | 22,158 | 1.26% |
| Illinois | IL | 12,716,164 | 2,807,983 | 22.08% | 2,949,760 | 23.20% | 612,796 | 4.82% |
| Indiana | IN | 6,696,893 | 926,319 | 13.83% | 731,067 | 10.92% | 165,155 | 2.47% |
| Iowa | IA | 3,150,011 | 347,203 | 11.02% | 269,234 | 8.55% | 62,685 | 1.99% |
| Kansas | KS | 2,912,619 | 452,019 | 15.52% | 310,393 | 10.66% | 47,108 | 1.62% |
| Kentucky | KY | 4,461,952 | 461,338 | 10.34% | 377,879 | 8.47% | 124,493 | 2.79% |
| Louisiana | LA | 4,664,616 | 455,010 | 9.75% | 334,750 | 7.18% | 75,619 | 1.62% |
| Maine | ME | 1,340,825 | 61,772 | 4.61% | 58,113 | 4.33% | 18,077 | 1.35% |
| Maryland | MD | 6,037,624 | 680,847 | 11.28% | 589,586 | 9.77% | 133,745 | 2.22% |
| Massachusetts | MA | 6,873,003 | 1,099,693 | 16.00% | 1,133,977 | 16.50% | 282,649 | 4.11% |
| Michigan | MI | 9,973,907 | 775,809 | 7.78% | 735,507 | 7.37% | 218,353 | 2.19% |
| Minnesota | MN | 5,600,166 | 762,266 | 13.61% | 560,400 | 10.01% | 99,099 | 1.77% |
| Mississippi | MS | 2,981,835 | 301,060 | 10.10% | 195,670 | 6.56% | 40,505 | 1.36% |
| Missouri | MO | 6,124,160 | 697,696 | 11.39% | 515,537 | 8.42% | 113,043 | 1.85% |
| Montana | MT | 1,061,705 | 126,116 | 11.88% | 87,962 | 8.29% | 17,093 | 1.61% |
| Nebraska | NE | 1,923,826 | 266,504 | 13.85% | 256,929 | 13.36% | 42,991 | 2.23% |
| Nevada | NV | 3,030,281 | 419,260 | 13.84% | 358,163 | 11.82% | 70,602 | 2.33% |
| New Hampshire | NH | 1,355,244 | 77,571 | 5.72% | 67,836 | 5.01% | 19,261 | 1.42% |
| New Jersey | NJ | 8,885,418 | 1,719,993 | 19.36% | 1,421,097 | 15.99% | 271,249 | 3.05% |
| New Mexico | NM | 2,097,021 | 178,753 | 8.52% | 128,770 | 6.14% | 42,965 | 2.05% |
| New York | NY | 19,514,849 | 3,698,438 | 18.95% | 4,581,391 | 23.48% | 692,324 | 3.55% |
| North Carolina | NC | 10,386,227 | 636,173 | 6.13% | 595,690 | 5.74% | 216,742 | 2.09% |
| North Dakota | ND | 760,394 | 110,813 | 14.57% | 69,887 | 9.19% | 11,204 | 1.47% |
| Ohio | OH | 11,675,275 | 1,512,456 | 12.95% | 1,142,243 | 9.78% | 234,274 | 2.01% |
| Oklahoma | OK | 3,949,342 | 460,858 | 11.67% | 315,979 | 8.00% | 57,939 | 1.47% |
| Oregon | OR | 4,176,346 | 580,397 | 13.90% | 491,015 | 11.76% | 131,661 | 3.15% |
| Pennsylvania | PA | 12,794,885 | 2,032,704 | 15.89% | 1,666,183 | 13.02% | 296,809 | 2.32% |
| Rhode Island | RI | 1,057,798 | 107,514 | 10.16% | 108,749 | 10.28% | 24,919 | 2.36% |
| South Carolina | SC | 5,091,517 | 421,412 | 8.28% | 320,628 | 6.30% | 77,393 | 1.52% |
| South Dakota | SD | 879,336 | 82,623 | 9.40% | 65,412 | 7.44% | 13,673 | 1.55% |
| Tennessee | TN | 6,772,268 | 566,253 | 8.36% | 495,610 | 7.32% | 125,322 | 1.85% |
| Texas | TX | 28,635,442 | 4,109,907 | 14.35% | 3,375,158 | 11.79% | 781,061 | 2.73% |
| Utah | UT | 3,151,239 | 427,757 | 13.57% | 327,323 | 10.39% | 98,253 | 3.12% |
| Vermont | VT | 624,340 | 25,665 | 4.11% | 24,833 | 3.98% | 5,233 | 0.84% |
| Virginia | VA | 8,509,358 | 980,822 | 11.53% | 773,988 | 9.10% | 207,411 | 2.44% |
| Washington | WA | 7,512,465 | 963,339 | 12.82% | 901,217 | 12.00% | 196,713 | 2.62% |
| West Virginia | WV | 1,807,426 | 101,675 | 5.63% | 79,158 | 4.38% | 22,586 | 1.25% |
| Wisconsin | WI | 5,806,975 | 612,177 | 10.54% | 501,934 | 8.64% | 127,510 | 2.20% |
| Wyoming | WY | 581,348 | 68,324 | 11.75% | 54,478 | 9.37% | 8,685 | 1.49% |
| Total |  | 326,569,308 | 43,822,211 | 13.42% | 39,191,398 | 12.00% | 8,879,860 | 2.72% |

**Table 1. (Continued).**
| State |  | Exposed 70-80 dB |  | Exposed 80-90 dB |  | Exposed 90+ dB |  |
| --- | --- | --- | --- | --- | --- | --- | --- |
| Alabama | AL | 22,289 | 0.46% | 3,021 | 0.06% | 262 | 0.01% |
| Alaska | AK | 2,349 | 0.32% | 56 | 0.01% | 4 | 0.00% |
| Arizona | AZ | 40,748 | 0.57% | 9,166 | 0.13% | 89 | 0.00% |
| Arkansas | AR | 11,045 | 0.37% | 2,165 | 0.07% | 167 | 0.01% |
| California | CA | 672,518 | 1.71% | 249,542 | 0.63% | 3,014 | 0.01% |
| Colorado | CO | 27,845 | 0.49% | 4,162 | 0.07% | 178 | 0.00% |
| Connecticut | CT | 23,507 | 0.66% | 3,498 | 0.10% | 237 | 0.01% |
| Delaware | DE | 5,867 | 0.61% | 375 | 0.04% | 47 | 0.00% |
| District of Columbia | DC | 5,229 | 0.74% | 364 | 0.05% | 61 | 0.01% |
| Florida | FL | 149,988 | 0.71% | 24,145 | 0.11% | 506 | 0.00% |
| Georgia | GA | 42,151 | 0.40% | 9,351 | 0.09% | 436 | 0.00% |
| Hawaii | HI | 11,944 | 0.84% | 855 | 0.06% | 3 | 0.00% |
| Idaho | ID | 4,568 | 0.26% | 510 | 0.03% | 30 | 0.00% |
| Illinois | IL | 99,178 | 0.78% | 16,629 | 0.13% | 2,847 | 0.02% |
| Indiana | IN | 34,426 | 0.51% | 5,284 | 0.08% | 545 | 0.01% |
| Iowa | IA | 16,768 | 0.53% | 2,285 | 0.07% | 242 | 0.01% |
| Kansas | KS | 16,158 | 0.55% | 1,320 | 0.05% | 252 | 0.01% |
| Kentucky | KY | 24,682 | 0.55% | 3,598 | 0.08% | 233 | 0.01% |
| Louisiana | LA | 19,955 | 0.43% | 1,245 | 0.03% | 170 | 0.00% |
| Maine | ME | 3,111 | 0.23% | 148 | 0.01% | 24 | 0.00% |
| Maryland | MD | 38,292 | 0.63% | 6,140 | 0.10% | 271 | 0.00% |
| Massachusetts | MA | 58,093 | 0.85% | 10,560 | 0.15% | 708 | 0.01% |
| Michigan | MI | 43,531 | 0.44% | 8,085 | 0.08% | 168 | 0.00% |
| Minnesota | MN | 35,130 | 0.63% | 2,813 | 0.05% | 400 | 0.01% |
| Mississippi | MS | 9,020 | 0.30% | 794 | 0.03% | 132 | 0.00% |
| Missouri | MO | 36,880 | 0.60% | 5,263 | 0.09% | 401 | 0.01% |
| Montana | MT | 2,844 | 0.27% | 177 | 0.02% | 38 | 0.00% |
| Nebraska | NE | 9,189 | 0.48% | 1,835 | 0.10% | 236 | 0.01% |
| Nevada | NV | 19,773 | 0.65% | 5,942 | 0.20% | 19 | 0.00% |
| New Hampshire | NH | 4,712 | 0.35% | 124 | 0.01% | 18 | 0.00% |
| New Jersey | NJ | 72,672 | 0.82% | 8,336 | 0.09% | 924 | 0.01% |
| New Mexico | NM | 6,768 | 0.32% | 1,399 | 0.07% | 42 | 0.00% |
| New York | NY | 191,507 | 0.98% | 46,899 | 0.24% | 2,277 | 0.01% |
| North Carolina | NC | 41,654 | 0.40% | 3,879 | 0.04% | 150 | 0.00% |
| North Dakota | ND | 2,894 | 0.38% | 153 | 0.02% | 48 | 0.01% |
| Ohio | OH | 68,199 | 0.58% | 9,965 | 0.09% | 1,129 | 0.01% |
| Oklahoma | OK | 18,672 | 0.47% | 1,970 | 0.05% | 245 | 0.01% |
| Oregon | OR | 23,035 | 0.55% | 2,815 | 0.07% | 241 | 0.01% |
| Pennsylvania | PA | 81,433 | 0.64% | 8,828 | 0.07% | 1,729 | 0.01% |
| Rhode Island | RI | 6,601 | 0.62% | 278 | 0.03% | 43 | 0.00% |
| South Carolina | SC | 18,937 | 0.37% | 1,402 | 0.03% | 144 | 0.00% |
| South Dakota | SD | 3,217 | 0.37% | 203 | 0.02% | 15 | 0.00% |
| Tennessee | TN | 25,511 | 0.38% | 4,691 | 0.07% | 154 | 0.00% |
| Texas | TX | 212,888 | 0.74% | 43,813 | 0.15% | 1,781 | 0.01% |
| Utah | UT | 20,427 | 0.65% | 5,504 | 0.17% | 202 | 0.01% |
| Vermont | VT | 1,011 | 0.16% | 30 | 0.00% | 6 | 0.00% |
| Virginia | VA | 53,360 | 0.63% | 7,071 | 0.08% | 574 | 0.01% |
| Washington | WA | 43,657 | 0.58% | 5,769 | 0.08% | 248 | 0.00% |
| West Virginia | WV | 6,559 | 0.36% | 647 | 0.04% | 51 | 0.00% |
| Wisconsin | WI | 30,739 | 0.53% | 3,110 | 0.05% | 245 | 0.00% |
| Wyoming | WY | 1,815 | 0.31% | 114 | 0.02% | 21 | 0.00% |

|  |  |  |  |  |  |  |  |
| --- | --- | --- | --- | --- | --- | --- | --- |
| Total |  | 2,423,347 | 0.74% | 536,327 | 0.16% | 22,007 | 0.01% |

**Figure 1.**
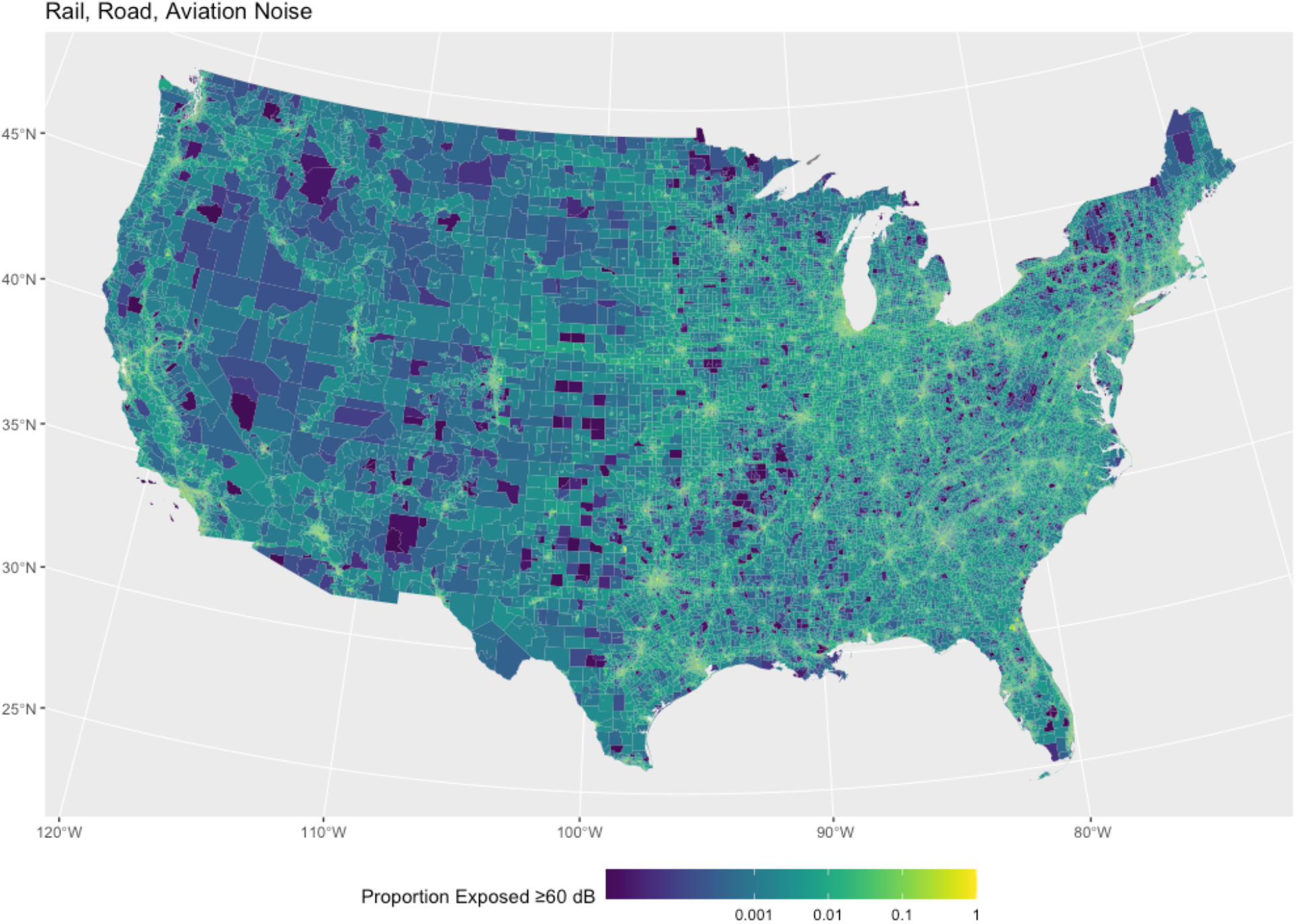
The National Transportation Noise Exposure Map for the continental United States in 2020.

In terms of exposure and health risk, California had the highest proportion of its population (6.5% of population) that was exposed to transportation noise levels ≥ 60 dB LAeq (Figure 2). Completing the top-five states, the District of Columbia and Illinois had the next largest proportions exposed both with 5.8%, Massachusetts with 5.1%, and New York with 4.8% of its population exposed.

**Figure 2.**
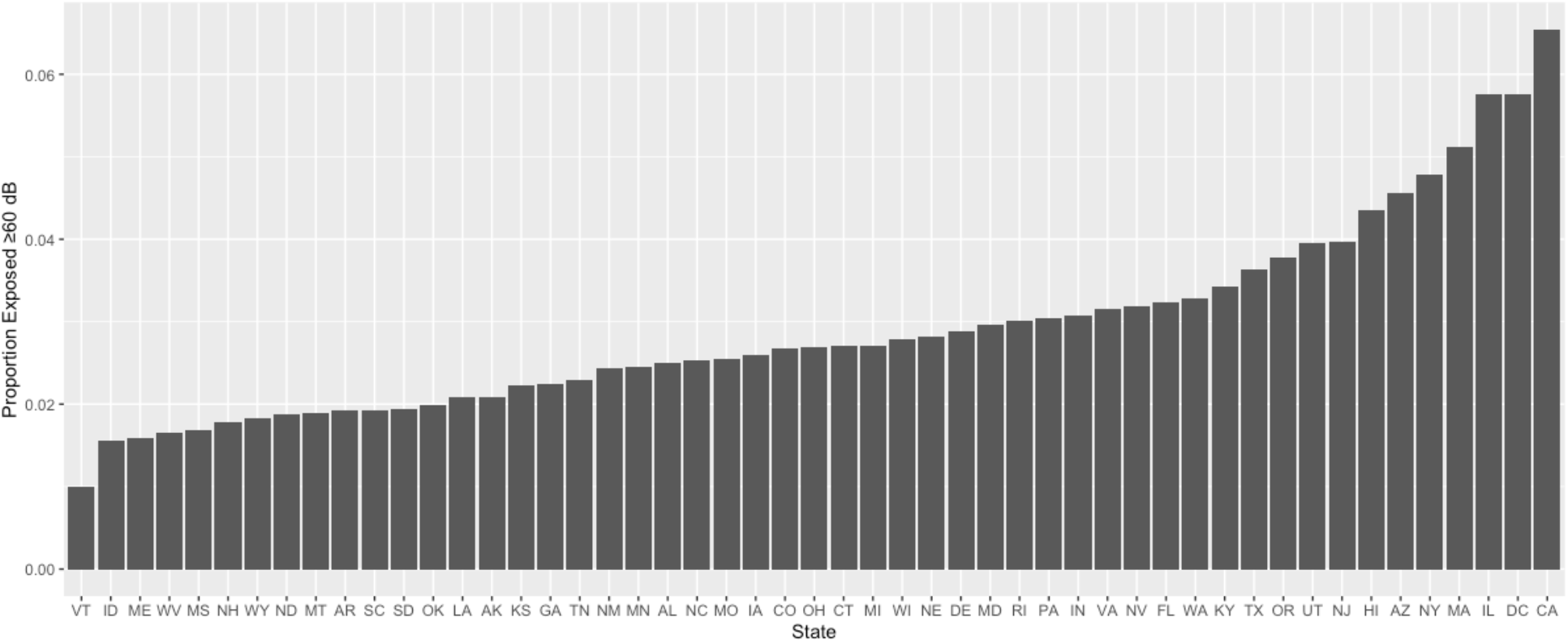
Ranking of the proportion of population with transportation noise exposures states ≥ 60 dB by state.

However, in terms of public health burden and the numbers of persons exposed to transportation noise, the more populated states tend to dominate the ranking (Figure 3). California had more than twice the number of people (approximately 2.6 million people) exposed to LAeq ≥ 60 dB transportation noise in 2020 than the next highest state, Texas with 1.0 million people exposed to these noise levels. The remaining top-five states in decreasing order of burden were New York, Illinois, and Florida.

**Figure 3.**
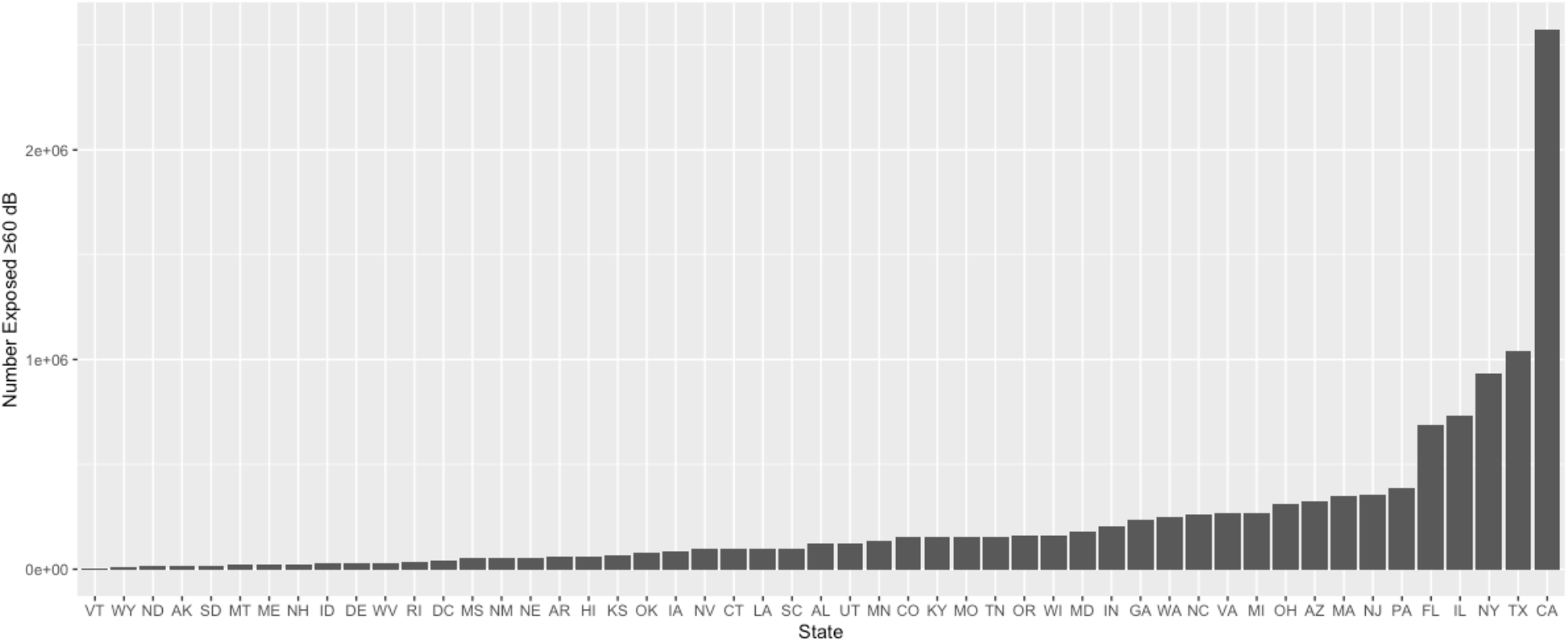
Ranking of the number of people with transportation noise exposures states ≥ 60 dB by state.

The spatial patterns of tract-level exposures largely follow the underlying source geographic distributions. Comparing maps of successively higher exposure categories in Figures 1, S3 and S4 for population proportions exposed to transportation noise levels ≥ 60, ≥ 70 and ≥ 80 dB LAeq, respectively reveals high noise exposures for those living along the network of highways and major roads across the U. S. As we zoom into specific regions, such as in the Seattle, King County region of Washington state, we can see the impact of airports on the proportion population exposed to noise. All three airports: Seattle-Tacoma International Airport, Boeing Field, and Renton Municipal Airport are located in the higher exposed regions (Figure S5), and the region most exposed to noise (in yellow color) is aligned with the north-south directions of the flight paths for Sea-Tac Airport.

Noise levels were found to vary considerably within census tracts due to the rapid noise attenuation with sound propagation distance. As an example, Figure S6 illustrates the distributions of sound levels for 30 m resolution pixels in the NTNM raster map for each census tract within King County, Washington. The boxplots are ordered by increasing median noise level in each tract, and yet large and small interquartile ranges can be observed through the range of median noise levels. The average interquartile range across the census tracts in this county is 7.2 dB LAeq. In contrast the average interquartile range across census block groups in the same county is less at 6.7 dB LAeq. This indicates that there can be considerable variation in noise levels within a census tract, and that there is less variation when using smaller census geographies such as the block group. In general, because of the variations in noise levels within either census tracts or block groups, the median may not be a good representation of noise exposure within these census boundaries. For this reason, we employed smaller block group level exposure assignments to compute numbers of exposed to separate noise level categories before aggregating these numbers to tract level estimates.

## Discussion

This paper presents methods used to develop the NTNE Map, a population exposure map for transportation-related noise at the census tract-level for the U. S. Our map focuses on transportation-related noise, which is a major contributor to noise in populated areas. This map benefits from methodological improvements over previous national noise exposure modeling approaches, notably in the use of the NTNM and the collective noise source data and modeling efforts of federal aviation, road, and rail agencies. These models utilize transportation scheduling and count data, noise emissions, and sound propagation principles. This differs from other studies that utilize spatial models fit to sound monitoring data (19–21), which depend on the availability and the generalizability of sampling locations for sound monitoring data. In some cases, entire states may not have appropriate sound monitoring data to fit spatial land use regression type sound models (see Figure 1 in (23)). Also, compared to previous studies that have used the BTS NTNM (35), our map provides some potential methodological improvements, including the computation of block group level population exposures before aggregation to the census tract, which allows for better accounting of uneven spatial distributions of population within census tracts, as well as the variations in noise that exist within tracts. Additionally, our methods retain the population proportions and numbers exposed to different noise level categories, which is useful for applying noise exposure-response functions in health impact assessments. Use zonal statistics such as the median non-missing noise level within a census tract computed in other studies (26) may not be appropriate for use in health impact assessments due to the large variation in noise within tract.

The previous EPA 1981 nationwide exposure assessment estimated that 23.4% of the U. S. population was exposed to Ldn ≥ 60 dB, 2.9 % was exposed to ≥ 70 dB, and 0.05 % was exposed to ≥ 80 dB (22). These population exposure percentages were based on the Ldn noise metric, which accounts for a 10 dB nighttime penalty. Based on our NTNE Map, 3.6 %, 0.9 %, and 0.2 % were exposed to those same noise level categories in 2020, but for the LAeq noise metric. Direct comparisons are difficult because the Ldn metric will be higher than a LAeq due to the nighttime penalty incorporated in the Ldn. Also, EPA’s assessment includes transportation related exposures, as well as other community noise sources and occupational noise exposure. Nevertheless, we notice that at the highest ≥ 80 dB category, we estimate a greater population proportion exposed than EPA’s 1981 assessment, which potentially highlights the disproportionately high levels of exposures for those that live closest to major transportation routes and hubs.

The disproportionate exposure to community noise has been previously studied by others, who have found that nighttime noise levels may be higher in urban neighborhoods with predominately black residents compared to neighborhoods with predominately white residents, and higher nighttime noise levels in neighborhoods with a larger percentage of the residents living below the federal poverty threshold (26). This previous work mentions potential explanations for such disparity in exposure, which may have to do with historical land use issues related to the types of noise sources (e.g., airplanes and roadway traffic) that exist in particular neighborhoods. This work also notes that ambient noise exposures may not fully represent household and personal exposures and health impacts, which are also affected by housing quality and noise mitigation measures, occupational noise exposure, and co-exposures such as to air pollution, and co-morbidities that may place individuals at greater risk for noise-related health issues.

Other researchers who have used the BTS transportation noise map have also found evidence of disparate noise exposures. A study that overlaid the noise map on the location of U. S. public schools found that high exposure was related to greater likelihood of students eligible for free/reduced meals, greater likelihood to be Hispanic, black, or Asian/Pacific Islander students (36). These authors have also used the transportation noise map to look more broadly at environmental justice issues, and have found that noise is associated with lower socioeconomic status generally, and affects greater proportions of Hispanic, black, Asian, Pacific Islander, and middle/working-aged groups (35). These studies illustrate the potential for a regularly updated and easily accessible national transportation exposure map, like NTNE Map, to be used in more studies to understand the context and causes for transportation noise exposure inequity.

Because the BTS produces regular updates to the NTNM for trend analyses by transportation agencies, there is an opportunity for regular updating of exposure estimates for our NTNE Map to assess trends in exposure over time. Currently, model results for the NTNM are available for the years 2016, 2018, and 2020. This may facilitate epidemiologic studies that require longitudinal noise exposures, as well as health impact assessments estimating potential changes in noise-related health outcomes over time due to changes in transportation.

There are potential limitations of the national exposure map that should be considered for exposure assessments in subsequent studies. First, the map only considers transportation-related noise. Thus, it is likely an underestimate of community noise exposures, which include exposures to other noise sources such as the soundscape of the natural environment, people noise, entertainment, construction, commercial and industrial, machinery, etc. Moreover, not all transportation-related sources are included in the noise models. Notably, military airport traffic is not included in the aviation noise model. Also, maritime traffic noise is not included in the model. Therefore, exposures are probably not accurate in the map for residents living near military airports or maritime ports and their traffic routes. This is an important limitation for environmental justice studies, as both military airports and port communities have loud noise sources that will not be well-represented on the map. Second, as a neighborhood tract level ambient transportation noise exposure map, it does not account for individual level factors that may affect exposure, such as housing conditions noise mitigation efforts, work exposure, mobility across varying noise micro environments, etc. Efforts are need to compare measured personal noise exposures to noise exposures assigned to their residential census tract location or time spent in different tracts due to personal mobility. Third, the mapped exposures are computed using the LAeq noise metric, yet many noise exposure-response functions are based on the Ldn or Lden noise metrics, which incorporate dB penalties during evening and nighttime hours. Therefore, use the LAeq from the noise map in some exposure-response functions will likely underestimate potential population health impacts. Fourth, although the individual noise models for aviation, roadway, and rail traffic have undergone some validation, there is still a need for validation studies to determine the relationships between modeled sum of transportation-related noise levels and real-world measured community noise levels.

Despite these limitations, the NTNM has been found to be useful in previous studies. With the availability of our new NTNE Map that characterizes population exposures at the census tract level, we hope that it will be easier for groups to integrate noise exposures into a variety of studies, including regional and national health impact assessments, environmental justice studies, and epidemiologic studies that consider noise as either primary exposures or co-exposures of interest.

## Data Availability

All data produced are available online at:
https://deohs.washington.edu/national-transportation-noise-exposure-map

