## Supplemental Information Figures for "The National Transportation Noise Exposure Map"

### Supplemental Info

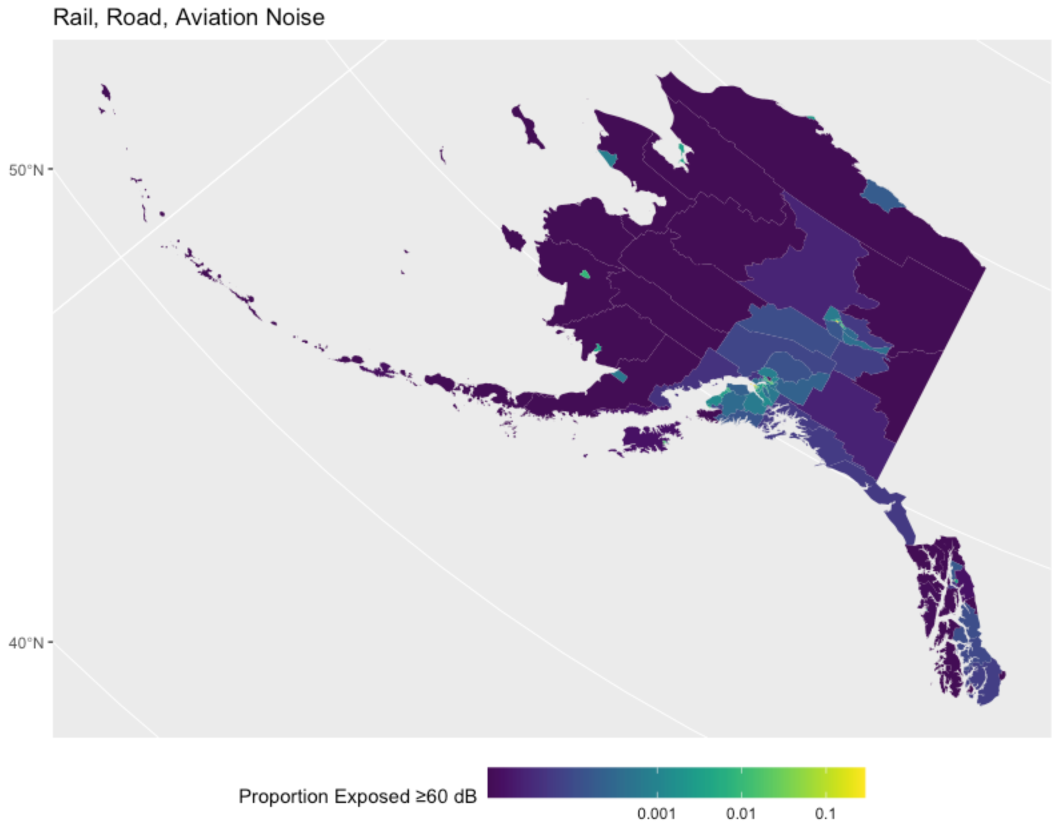


**Figure S1. The National Transportation Noise Exposure Map for Alaska in 2020.**

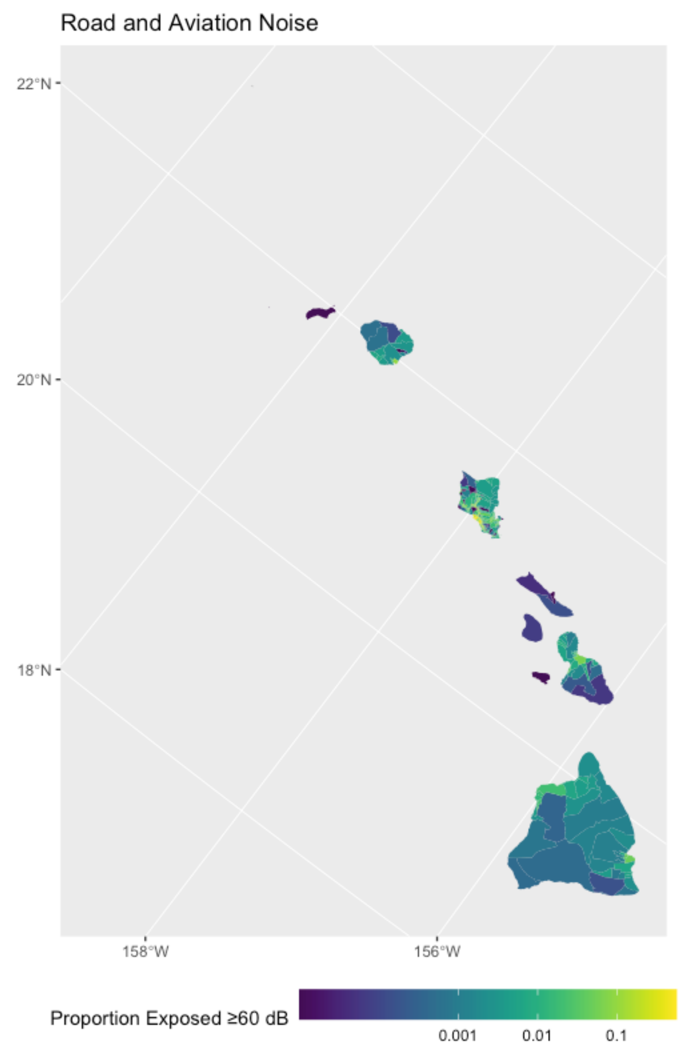


**Figure S2. The National Transportation Noise Exposure Map for Hawaii in 2020.**

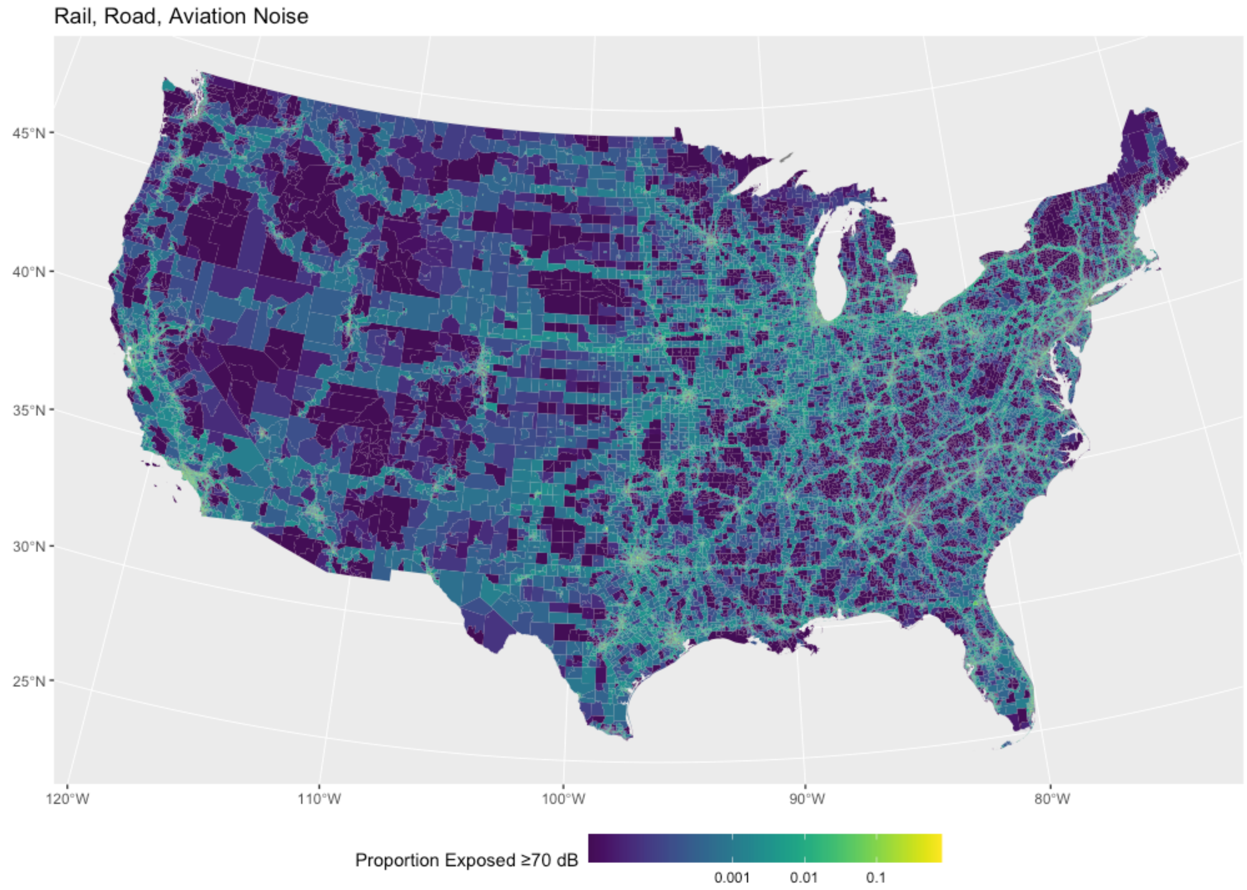


**Figure S3. The National Transportation Noise Exposure Map for exposures ≥ 70 dB**

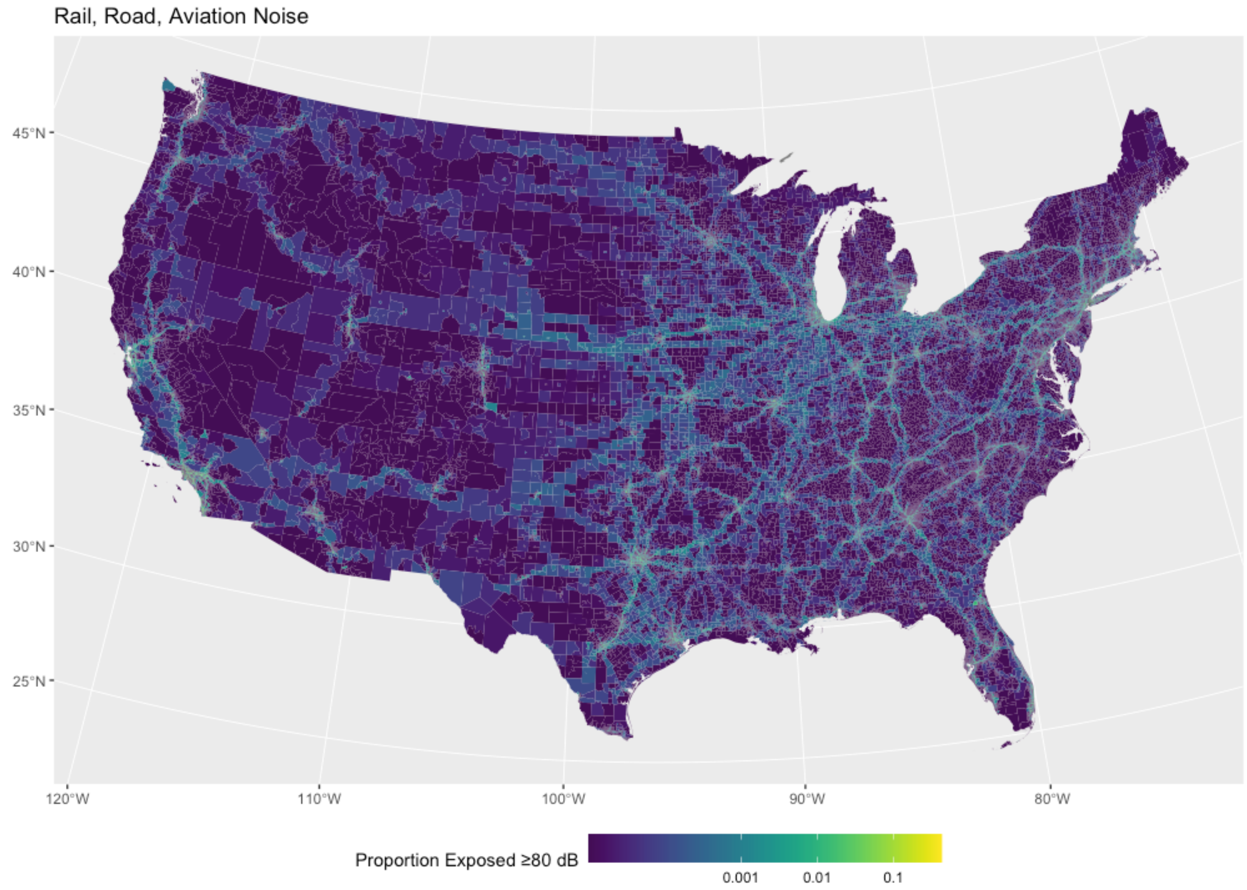


**Figure S4. The National Transportation Noise Exposure Map for exposures ≥ 80 dB**

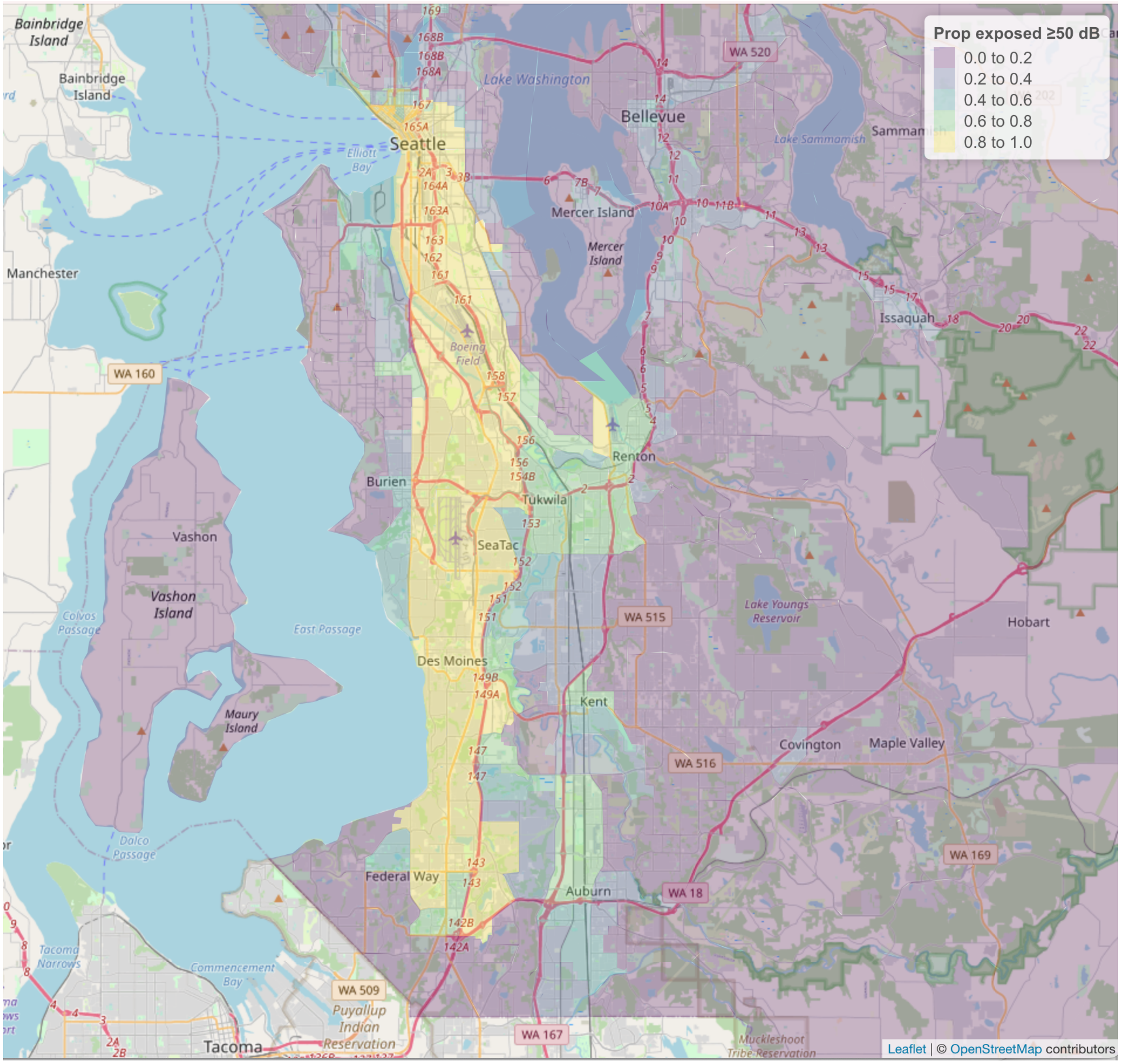


**Figure S5. Seattle King County, Washington transportation noise exposures ≥ 50 dB. (Underlying basemap from OpenStreetMap)**

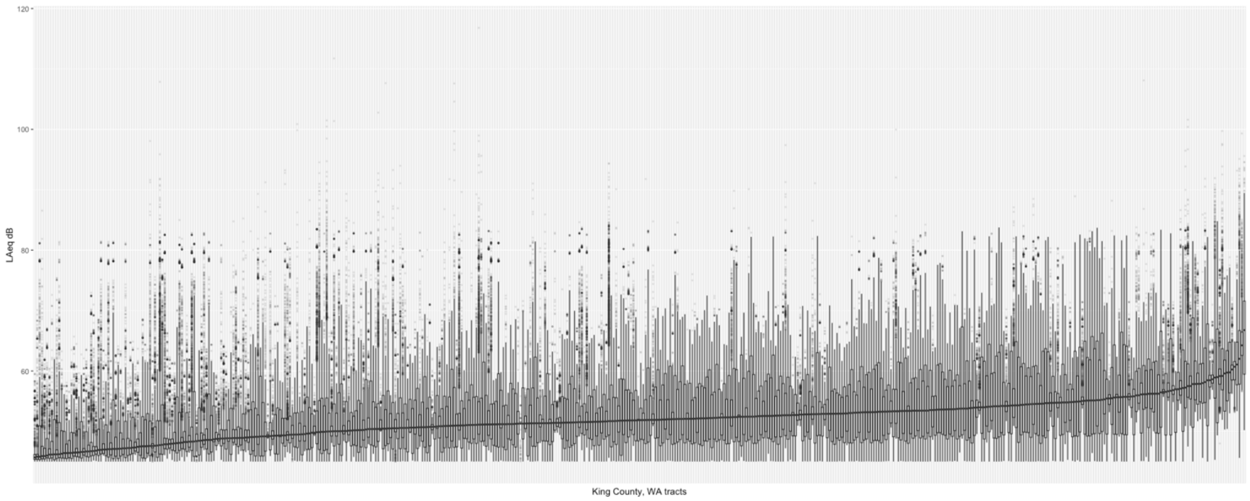


**Figure S6. Boxplots of the distribution of 30 m resolution noise level pixels in census tracts in Seattle King County, Washington**
